# A New Hope for Liquid Biopsies: Early Detection of Pancreatic Cancer By Means of Protease Activity Detection in Serum Applying a Hierarchical Decision Structure

**DOI:** 10.1101/2022.10.18.22281240

**Authors:** Obdulia Covarrubias-Zambrano, Deepesh Agarwal, Madumali Kalubowilage, Sumia Ehsan, Asanka S. Yapa, Jose Covarrubias, Anup Kasi, Balasubramaniam Natarajan, Stefan H. Bossmann

**Author notes:** Obdulia Covarrubias-Zambrano and Deepesh Agarwal share first authorship. Corresponding authors: Prof. Dr. Stefan H. Bossmann, University of Kansas Medical Center, Department of Cancer Biology, Wahl Hall West, 2001, Mailstop 1071, 3901 Rainbow Boulevard, Kansas City, KS 66160, Prof. Dr. Balasubramaniam Natarajan, Kansas State University, Department of Electrical and Computer Engineering, Manhattan, KS 66506; Associate Prof. Anup Kasi, MD, MPH, University of Kansas Cancer Center, 2330 Shawnee Mission Pkwy Ste 210, Westwood KS, 66205.

## Abstract

Over the last 6 years, five-year survival rate for pancreatic cancer patients has increased from 6 to 10% after the initial diagnosis, which makes it one of the deadliest cancer types. This disease is known as the “silent killer” because early detection is challenging due to the location of the pancreas in the body and the nonspecific clinical symptoms. The Bossmann group has developed ultrasensitive nanobiosensors for protease/arginase detection comprised of Fe/Fe_3_O_4_ nanoparticles, cyanine 5.5, and designer peptide sequences linked to TCPP. Initial data obtained from both gene expression analysis and protease/arginase activity detection in serum indicated the feasibility of early pancreatic cancer detection. Several matrix metalloproteinases (MMPs, -1, -3, and -9), cathepsins (CTS) B and E, neutrophil elastase, and urokinase plaminogen activator (uPA) have been identified as candidates for proximal biomarkers. In this study, we have confirmed our initial results from 2018 performing serum sample analysis assays using a larger group sample size (n=159), which included localized (n=33) and metastatic pancreatic cancer (n=50), pancreatitis (n=26), and an age-matched healthy control group (n=50). The data obtained from the eight nanobiosensors capable of ultrasensitive protease and arginase activity measurements were analyzed by means of an optimized information fusion-based hierarchical decision structure. This permits the modeling of early-stage detection of pancreatic cancer as a multi-class classification problem. The most striking result is that this methodology permits the detection of localized pancreatic cancers from serum analyses with 96% accuracy.

## INTRODUCTION

Pancreatic cancer (PC) is the third deadliest cancer type for men and women, but it is expected to become the second leading cause of cancer-related death in the US by 2030^1^. Five-year survival rate has increased from 6 to 10% within the last 6 years, being the lowest 5-year survival rate; however, PC is one of the few cancer types for which survival has not improved significantly in the last 40 years^2^. PC is frequently misdiagnosed, due to the absence of early warning signs or misleading symptoms, rapid progression, and resistance to drug treatments, and for this reason it is known as the “silent killer” ^1-3^. PC is diagnosed after it has metastasized and is inoperable in approximately 80-85% of patients, and for patients with localized tumors detected, prognosis remains poor, with a lower than 20% patients surviving 5 years after tumor removal^3^. Therefore, a feasible and cost-effective detection method capable of detecting PC at the localized stage would be of great value, preferably one that works by means of a simple blood test, better known as liquid biopsy^4^.

Liquid biopsy or fluid phase biopsy consists of the sampling and analysis of tumor materials or molecules found in bodily fluids, such as blood, urine, or saliva. Compared to a solid tissue biopsy, liquid biopsy is less invasive and easily repeatable since bodily fluids are readily accessible^5^. Currently, two different methods of liquid biopsy are explored in the clinic, Circulating Tumor Cells (CTC), composed of singular units or clusters of cancer cells that split away from primary tumor, and Cell-free DNA (cfDNA) composed of nucleic caid fragments released into the bloodstream^6^. Although, these liquid biopsies are able to identify advanced solid tumors, they are less effective when it comes to early-stage tumors^7-9^. With the exception of the protease-activity technology discussed in this manuscript, none of the “classic” approaches to liquid biopsies, such as capture and detection of circulating tumor cell or circulating tumor DNA, DNA-methylation studies, or the analysis of the content of extracellular vesicles are capable of detecting early-stage of PC in a reliable manner^10-12^.

Since 2007, established panels of proteases and arginase activities in serum have been measured using this technology^13-18^. More specifically, a panel of seven proteases (caspases B and E, matrix metalloproteinases (MMPs) 1, 3, and 9, urokinase plasminogen activator (UpA), neutrophil elastase and arginase were established by the Bossmann group for early detection of PDAC in 2018^13^. This current manuscript is a continuation study from this previously developed panel, which was selected utilizing data from NCBI Gene Expression Omnibus dataset^19,20^. One of the advantages of Fe/Fe_3_O_4_-based nanobiosensor technology for protease/arginase detection is that the materials cost for a panel of eight enzymes for early pancreatic cancer detection from serum is currently less than 10 dollars. This will abet the world-wide utilization of this technology.

Function principles for this nanobiosensor technology are shown in Figure 1. Nanobiosensors consist of a Fe/Fe_3_O_4_ core/shell nanoparticle (diameter 15 nm) coated with dopamine, this core nanoparticle is tethered to an oligopeptide labeled with the tetrakis-carboxyphenyl-porphyrin (TCPP) fluorophore and to the FRET acceptor cyanine 5.5 (Cy5.5) by a stable amide bond.

**Figure 1.**
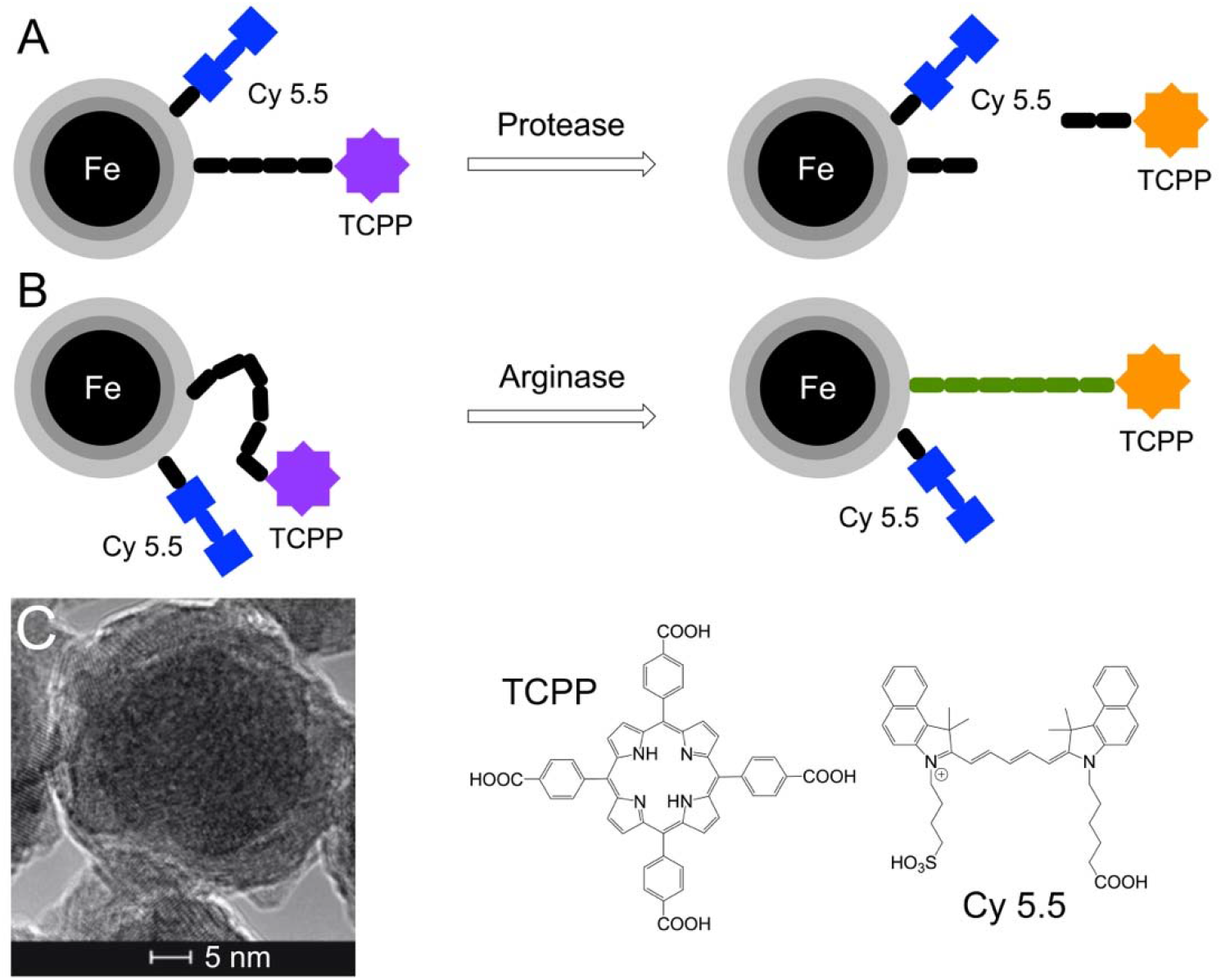
**A**: Design of a Fe/Fe_3_O_4_-nanoparticle based nanobiosensor for protease detection. Gray: Fe_3_O_4_; light gray: dopamine. In the “off-state”, the fluorophore TCPP (tetrakis-carboxyphenyl-porphyrin) is quenched by Cy 5.5 (cyanine 5.5) by a FRET mechanism^22^ and by the Fe(0) core by means of plasmonic quenching^14^. B: Design of a Fe/Fe_3_O_4_-nanoparticle based nanobiosensor for arginase detection. Due to posttranslational enzymatic modification (arginase converts arginine to ornithine within the oligopeptide^16^), the tether dynamics decrease and, consequently, the distance between TCPP and center-nanoparticle increases. This increase leads to the switch on of TCPP fluorescence; C: HRTEM of dopamine-coated Fe/Fe_3_O_4_ core/shell nanoparticles. The inner core (d=13 *±* 1.0 nm) consists of disordered Fe(0) crystallites^14^. The inorganic shell (d=2 *±* 0.5 nm) is comprised of Fe_3_O_4_. Dopamine is tightly bound to the surface of Fe_3_O_4_^25^, thus enhancing the water-dispersibility of the protected Fe/Fe_3_O_4_ nanoparticles.

Previous mathematical modeling demonstrated that 35 TCPP and 50 cyanine 5.5 units are bonded to one Fe/Fe_3_O_4_ center in average assuming a Poisson distribution^14,21^. Activity of these protease nanobiosensors consist of the proteolytic cleavage of the oligopeptide when each specific protease is present. This cleavage allows TCPP to escape both FRET and plasmonic quenching^14,22^, to trigger an increase in fluorescence signal which is detected by a standard clinical plate reader. The arginase nanobiosensor activity is not cleaved. Here, arginase performs a “post-translational” modification by converting peptide-bound arginine into ornithine^16^.

Ornithine changes the dynamic of the peptide tether, which increases the average distance between nanoparticle and TCPP^16^ and, consequently, TCPP fluorescence^22^. In table 1, peptides 1 to 7 have been optimized to be specifically cleaved by the protease they were designed for, while peptide 8 is post-translationally modified by arginase. Studies have demonstrated that the human degradome consists of 553 proteases and homologues discovered to date (21 aspartic, 143 cysteine, 186 metallo, 176 serine and 27 threonine proteases)^23^, and virtually all proteases form an interdependent proteolytic network^24^. Consequently, numerous active proteases present in serum could degrade peptides 1-8, with significantly slower kinetics. Therefore, a comparison between the selected panel of enzymes in serum of pancreatic cancer patients and healthy control group is required to detect cancer at the localized stage.

**Table 1:**
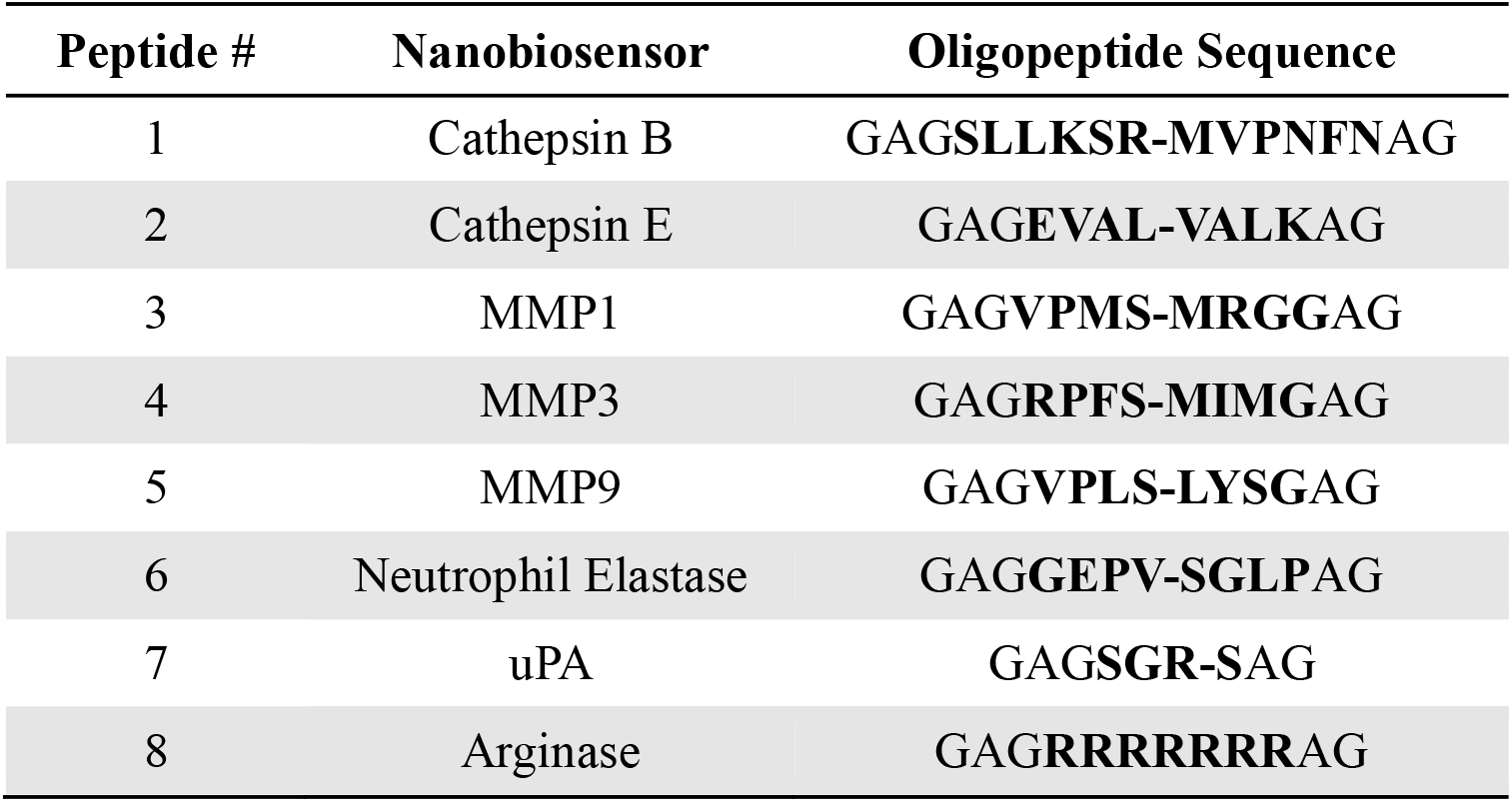
Peptides designed for proteases and arginase used to assemble nanobiosensors^13^. Each peptide was designed to preferentially react with its respective enzyme. However, serum contains multiple proteases that co-react with each peptide as well. Therefore, the peptide sequences have been optimized in our earlier work to react significantly faster with their “target enzyme” than with other enzymes that are present in serum ^13,15,16^.

| Peptide # | Nanobiosensor | Oligopeptide Sequence |
| --- | --- | --- |
| 1 | Cathepsin B | GAGSLLKSR-MVPNFNAG |
| 2 | Cathepsin E | GAGEVAL-VALKAG |
| 3 | MMP1 | GAGVPMS-MRGGAG |
| 4 | MMP3 | GAGRPFS-MIMGAG |
| 5 | MMP9 | GAGVPLS-LYSGAG |
| 6 | Neutrophil Elastase | GAGGEPV-SGLPAG |
| 7 | uPA | GAGSGR-SAG |
| 8 | Arginase | GAGRRRRRRRAG |

The statistically significant differences in protease/arginase activity pattern of the four groups, i.e., localized pancreatic cancer (n=33), metastatic pancreatic cancer (n=50), pancreatitis (n=26), and an age-matched healthy control group (n=50), are established by harnessing these Fe/Fe_3_O_4_-based nanobiosensors. This is achieved by using elementary biostatistical tools like performing Welch tests^26^ and computing p-values between all the data groups^26,27^. However, the sample sizes of the examined groups are very small, thereby insufficient to investigate the broader feasibility of detecting pancreatic cancer at early stages. Moreover, it is desirable to extract maximal discriminatory power by protease/arginase information fusion rather than relying on basic statistical analyses. Therefore, we propose two information fusion-based decision frameworks for early detection of pancreatic cancer. Specifically, a hard hierarchical decision structure (HDS) paired with step-by-step feature engineering approach is introduced and shown to outperform conventional multi-class classification techniques. Additionally, a soft hierarchical decision structure (SDS) that provides confidence in anticipated decisions via probability values for each class is introduced. The primary goal of employing computational approaches for early pancreatic cancer detection is to detect the onset of pancreatic cancer in individuals with risk factors, such as smoking, heavy alcohol use, obesity, long-standing diabetes, and chronic pancreatitis, allowing for the maximum amount of time available for successful treatment with other modalities such as immunotherapy^28^.

## METHODS

### Synthesis of Nanobiosensors

The detail procedures used for synthesis of nanobiosensors and individual components (dopamine coated Fe/Fe_3_O_4_ nanoparticles, peptides, and both fluorescent dyes, TCPP, and cyanine 5.5) can be found in previous publications from the Bossmann research group^4,13-18^. After synthesizing all components, each nanobiosensor was assembled. Briefly, a first solution was prepared by dissolving 64 mg of protease sequence labeled with TCPP, 37 mg of cy5.5, 45 mg of EDC, and 45 mg of DMAP in 30 mL of anhydrous DMF. In a separate vial, a second solution was prepared by dispersing 450 mg of dopamine coated Fe/Fe_3_O_4_ nanoparticles in 10 mL of anhydrous DMF. Each solution vial was mixed and sonicated for 20 min. Then, both solutions were combined, sonicated for 10 more minutes, and incubated in a shaker at room temperature (298K) overnight. Following overnight incubation, the nanobiosensor was washed with anhydrous DMF (25 mL) four times and with cold diethyl ether (25 mL, 263 K) four times for removal of excess dye and unreacted material, which remained in solution. The nanobiosensor was collected by means of centrifugation between washes (5 min. at 10,000 RPM). Finally, the nanobiosensor was collected and dried using argon gas and kept at 253K under argon.

### Ethical Statement

This study was funded by American Cancer Society grant (IRG-16-194-07), awarded to the University of Kansas Medical Center (PI: Anup Kasi). Ethical approval for this study was obtained from the University of Kansas Medical Center (Reviewing IRB: IRB00000161, IRB#5929.

### Serum Samples Information

All serum samples were obtained from the Biospecimen Repository Facility of the University of Kansas Cancer Center^29^. The total sample number received and tested was n=159, divided into the following groups: localized pancreatic cancer (LPC) n=33, metastatic pancreatic cancer (MPC) n=50, pancreatitis n=26, and apparently healthy volunteers (control) n=50.

### Protease Activity in Serum Sample Measurements using Fluorescent Plate Reader

The procedure and information used for measurement of protease in serum sample was fully described in previous study published in 2018^13^. Briefly, a 25 μmol HEPES buffer (2-[4-(2-hydroxyethyl) piperazin-1-yl]ethanesulfonic acid) solution was enriched with 10 μmol of each cation, Ca(II), Mg(II), and Zn(II) at 298K and pH adjusted to 7.2 in order to ensure enzymatic activity on MMPs. Briefly, to maintain enzymatic activity, a first solution containing 25 μmol/L HEPES buffer was enriched with 10 μmol/L of Ca^2+^, Mg^2+^, Zn^2+^ with a pH of 7.2 at 298K. A second solution was then prepared for each nanobiobiosensor analyzed by completely dispersing 0.30 mg of nanobiosensor in 1 mL of HEPES buffer enriched with cations after sonicating solution for 15 min at room temperature (298K). After these two solutions were prepared, the following four sample solutions were prepared for the actual analysis part: *1-Blank* (130 μL HEPES buffer enriched with cations), *2-Sample Control* (125 μL HEPES buffer enriched with cations + 5 μL serum samples), *3-Assay Control* (125 μL nanobiosensor assay solution + 5 μL HEPES buffer enriched with cations), and *4-Assay* (125 μL nanobiosensor assay solution + 5 μL serum samples). From each sample solution, a volume of 130 μL was loaded into a 96-well plate (black, flat bottom), plating three replicates for each assay and serum samples (sample solution 4). After loading all samples onto the plate, it was incubated at 37°C for 60 minutes. After incubation time, fluorescent intensity was measured using a BioTek Synergy 2 plate reader, where both endpoint (421 nm excitation & 650 nm emission) and spectra scan (600-700 nm emission) data was collected.

### Statistical Analysis

Equal or unequal variance of raw data was determined using an F-test. Once variance was determined for each comparison data set, a t-test with equal or unequal variance (Welch’s t-test) was then performed to compare data within each group for each biomarker^26^. Significant differences between two groups were reported when the p-value was ≤ 0.05, and borderline significant differences between two groups were reported when the p-value was between 0.05 and 0.1. These significant differences were annotated on each bar graph.

### Information Fusion-based Decision Framework

This work models the problem of early-stage pancreatic cancer detection as a multi-class classification task. The data generated from the experiments belongs to one of the four classes, namely, “Healthy”, “Pancreatitis”, “Localized” pancreatic cancer and “Metastatic” pancreatic cancer. We propose two information fusion-based decision frameworks: (i) an HDS coupled with stepwise feature engineering technique for improved performance in comparison to traditional classification methodologies, and (ii) an SDS that conveys confidence in predicted labels to improve explanations and allow clinicians to make informed judgements.

#### 1. Hard Hierarchical Decision Structure

The central idea of the proposed information fusion-based HDS is that statistically most significant features are appropriately weighted at each hierarchical level to conduct an efficient binary classification task. The proposed HDS is presented in Figure 2. The following subsections elaborate the components used to construct the HDS.

**Figure 2.**
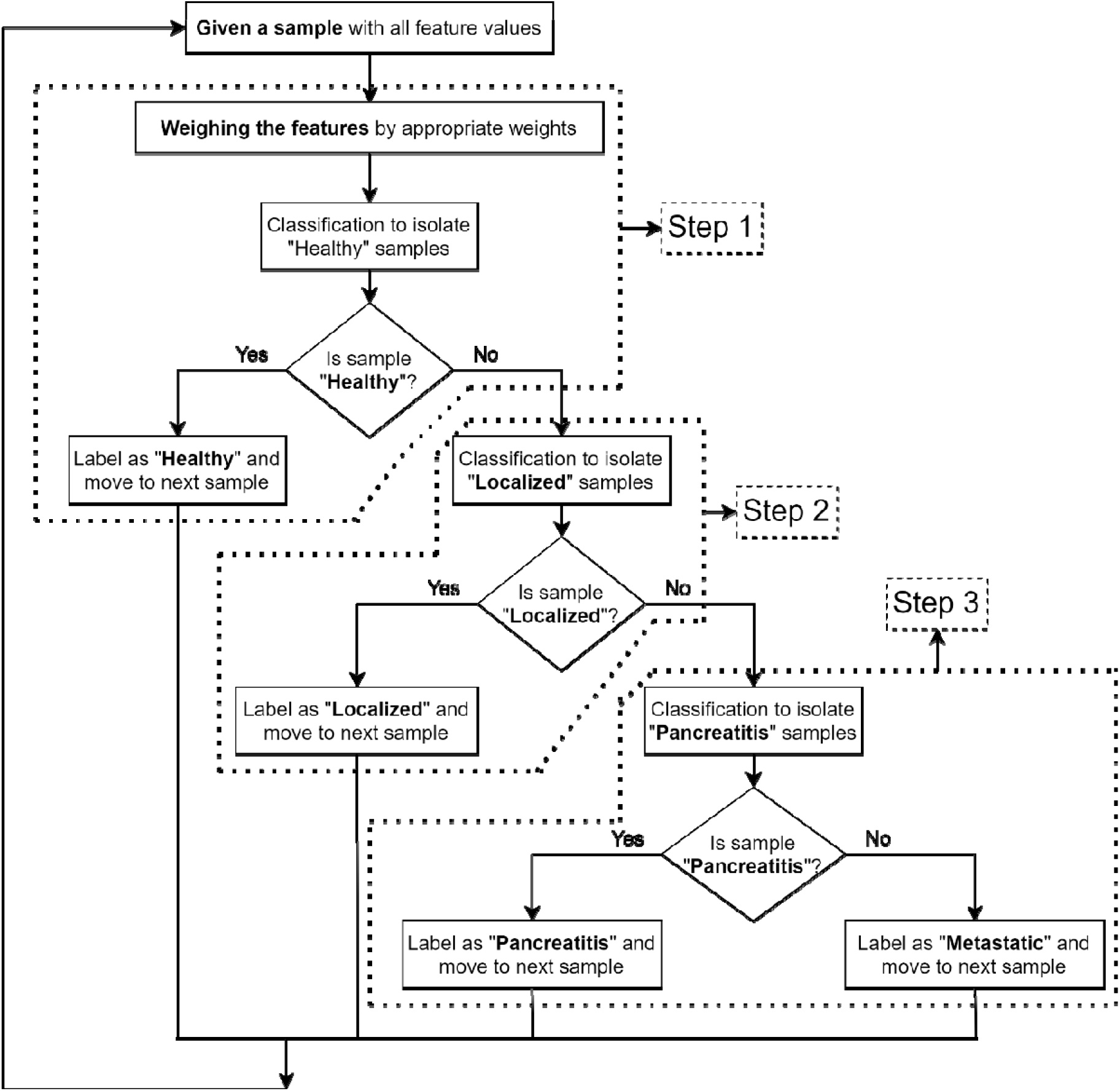
Proposed information fusion based HDS framework.

##### Calculating feature weights

The proposed HDS paradigm begins by determining whether the supplied sample is from the healthy (null hypothesis) or unhealthy (alternative hypothesis) group. In order to generate the corresponding binary classifier, feature engineering entails a suitable weighting of all features depending on their relative significance. The p-values of two-sample t-tests for all features across the set of measurements gathered from “healthy” and “non-healthy” groups are used to calculate these weights. Here, the null hypothesis is that the measurements in “healthy” and “non-healthy” groups belong to independent random samples from normal distributions with equal means. The corresponding test decision values and p-values are examined for all the features. The p-values were observed to be dispersed over a wide range and have a highly skewed distribution. As a result, the probability operations associated with them might generate extremely small values that are difficult to represent precisely. This leads to numerical errors like underflow or overflow.

In order to eliminate challenges concerning precision, the p-values are converted to a logarithmic scale for easier interpretation and analysis^30-32^. The negative values of natural logarithm of p-values, − log_*e*_*p* is computed for all the features and scaled, as shown in equation (1), to obtain the corresponding feature weights as

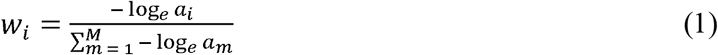

Here, *M* is the total number of features, *w*_*i*_ depicts the weight for feature *a* _*i*_ and − log_*e*_ *a* _*i*_ represents the negative value of natural logarithm of p-value corresponding to the feature *a*_*i*_. Table 2 demonstrates the p-values and computation of weights for all the features in the dataset under consideration.

**Table 2:**
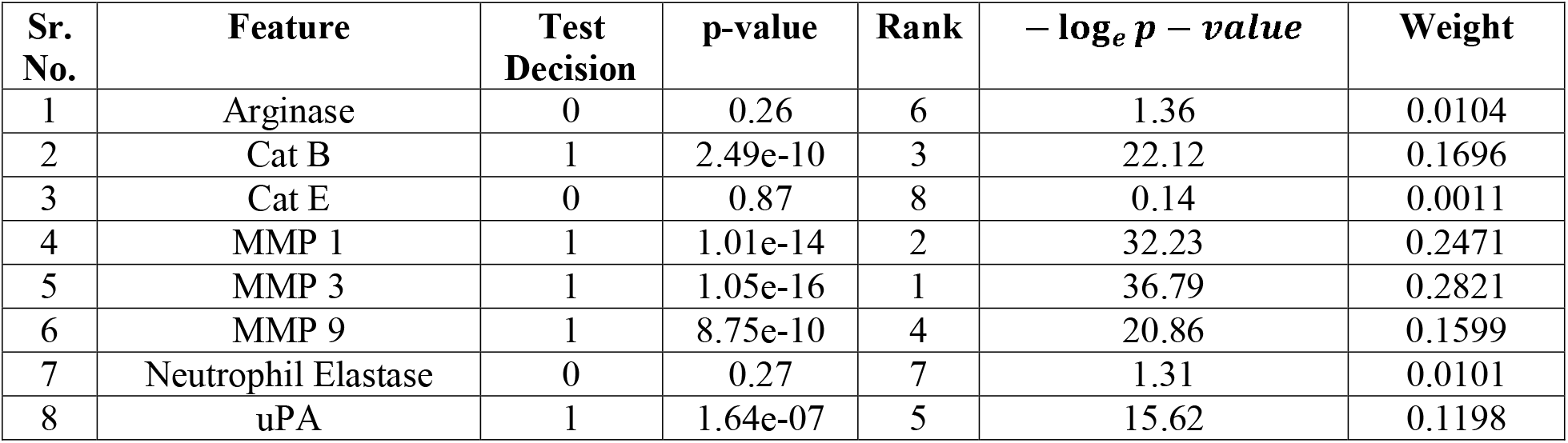
Calculating features weights. A test decision value of 0 indicates a failure to reject the null hypothesis at 95% confidence level: a value of 1 indicates rejection of the null hypothesis at 95% confidence level; a rank of 1 indicates that the corresponding feature is most important and that of 8 indicates that the corresponding feature is least important.

| Sr. No. | Feature | Test Decision | p-value | Rank | $-\log_e p - value$ | Weight |
| --- | --- | --- | --- | --- | --- | --- |
| 1 | Arginase | 0 | 0.26 | 6 | 1.36 | 0.0104 |
| 2 | Cat B | 1 | 2.49e-10 | 3 | 22.12 | 0.1696 |
| 3 | Cat E | 0 | 0.87 | 8 | 0.14 | 0.0011 |
| 4 | MMP 1 | 1 | 1.01e-14 | 2 | 32.23 | 0.2471 |
| 5 | MMP 3 | 1 | 1.05e-16 | 1 | 36.79 | 0.2821 |
| 6 | MMP 9 | 1 | 8.75e-10 | 4 | 20.86 | 0.1599 |
| 7 | Neutrophil Elastase | 0 | 0.27 | 7 | 1.31 | 0.0101 |
| 8 | uPA | 1 | 1.64e-07 | 5 | 15.62 | 0.1198 |

##### Feature selection for individual hierarchical levels

If a given sample is identified as “non-healthy” in the first hierarchical step, the next two steps are aimed at determining the type of abnormality involved. The second hierarchical step evaluates whether the given sample belongs to a “localized” group. If it is further recognized as “non-localized”, the last hierarchical level determines whether the sample belongs to “pancreatitis” or “metastatic” group. Unlike the binary classifier in the first hierarchical step, the ones in the second and third steps use a subset of features rather than using all the features obtained from the experiments. This process of feature engineering uncovers the most crucial features, simplifies the decision models, and fosters better interpretability. Further, this allows for shorter training times and less overfitting. The relevant features are identified by conducting a series of two-sample t-tests (as in the first hierarchical step) for all possible pairs of the class labels. The common features exhibiting the lowest p-values in all the relevant pairs of hypothesis tests are selected as admissible features for the corresponding binary classifier. For instance, a set of common features exhibiting lowest p-values in both the hypothesis tests, localized vs. pancreatitis, and localized vs. metastatic are selected as admissible features for the binary classifier in the second hierarchical step.

For the dataset under consideration, Cat B, MMP 1, Neutrophil Elastase and UpA are selected as features for binary classifiers in the second hierarchical step, and Arginase, MMP 1 and MMP 9 are the selected features for the third hierarchical step.

##### Training set

In the first hierarchical level, 80% of all instances in the dataset are selected randomly to train the binary classifier. The samples from the “healthy” group are isolated as a result of this step. So, 80% of the instances from “localized”, “pancreatitis” and “metastatic” groups are selected at random to form the training set for the binary classifier in the second hierarchical level. This step isolates the samples from “localized” group. Finally, 80% of the instances from the remaining two groups, i.e., “pancreatitis” and “metastatic” are randomly selected to train the binary classifier in the third hierarchical level. Such a strategy for building training sets for individual binary classifiers is highly beneficial when sample size of the dataset is limited.

#### 2. Soft Hierarchical Decision Structure

A four-class classifier offered by HDS does not incorporate any information pertaining to the confidence level associated with the decisions at each step. The proposed SDS framework addresses this limitation by providing confidences associated with the predicted labels in the form of probability values for each class. The proposed SDS is shown in Figure 3. It is essentially an extension of the HDS, where the prediction for each sample is accompanied with the probability values of that sample being affiliated to each of the four classes. The differences between these probability values indicate the degree of confidence associated with the predictions. For a given sample, if the probability value corresponding to one of the classes is significantly higher than others, the confidence associated with such a prediction is inferred to be HIGH. On the other hand, if the difference between the probability values corresponding to all the classes is not significant, the confidence in such a prediction would be LOW. This framework assists clinicians in determining whether more tests are necessary for correct diagnosis.

**Figure 3.**
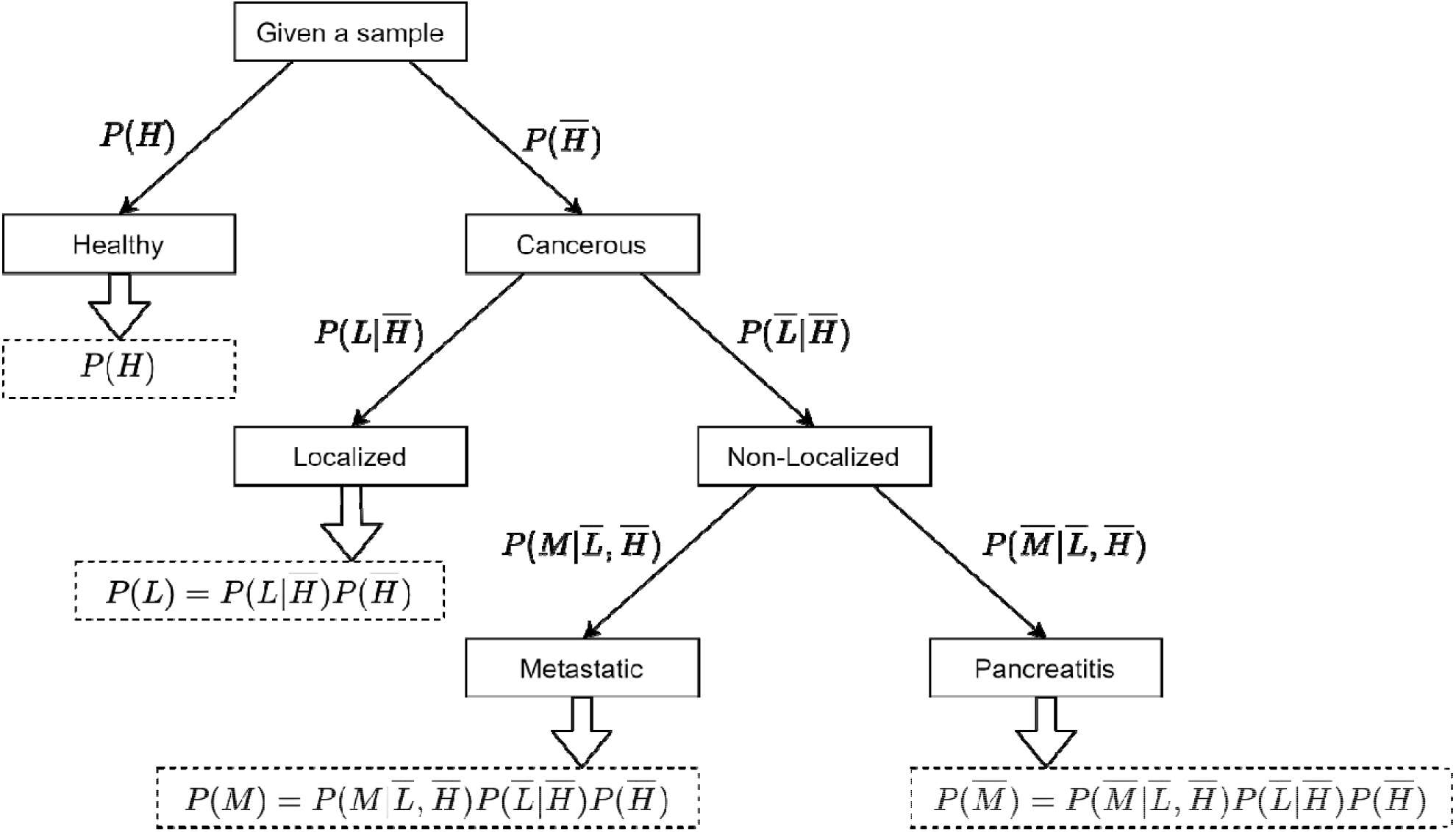
Proposed information fusion based SDS framework.

All steps in the SDS are probabilistic extensions of the HDS. For example, the first level in SDS results in two values indicating probabilities of the given sample being “healthy” or “non-healthy”, represented by and respectively. The second hierarchical level determines the probabilities of the given sample being “localized” or “non-localized”, given the condition that it belongs to “non-healthy” group, represented by and respectively. As a result, the probabilities of a sample being “localized” is calculated based on equation (2).

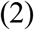

The last step in SDS computes the probabilities of the given sample being “metastatic” or “pancreatitis”, given the condition that it is “non-healthy” and “non-localized”, represented by and respectively. Finally, the probabilities of a sample being “metastatic” or “pancreatitis” is evaluated based on equations (3) and (4) respectively.

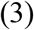

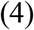

## RESULTS

### Quantified protease activity in serum samples

In our previous pancreatic cancer detection study published in 2018, Gene Expression Analysis (GEO) was used to identify and select protease candidates, there detailed information for selection criteria and a summary for gene regulation can be found, as well as a full description of synthesis, validation, and calibration for each nanobiosensor used in this study^14^. All eight nanobiosensors (arginase, cathepsin B, cathepsin E, MMP1, MMP3, MMP9, neutrophil elastase, and uPA) were used to measure enzymatic activity in serum samples from four different disease groups: localized pancreatic cancer, metastatic pancreatic cancer, pancreatitis, and a ‘healthy’ control. Figure 4 to 11 compare the enzymatic activity for the disease group for each nanobiosensor tested. Figure 4 demonstrated that the enzymatic activity of arginase was significantly upregulated in serum from the metastatic and control group compared to both localized and pancreatitis groups.

**Figure 4.**
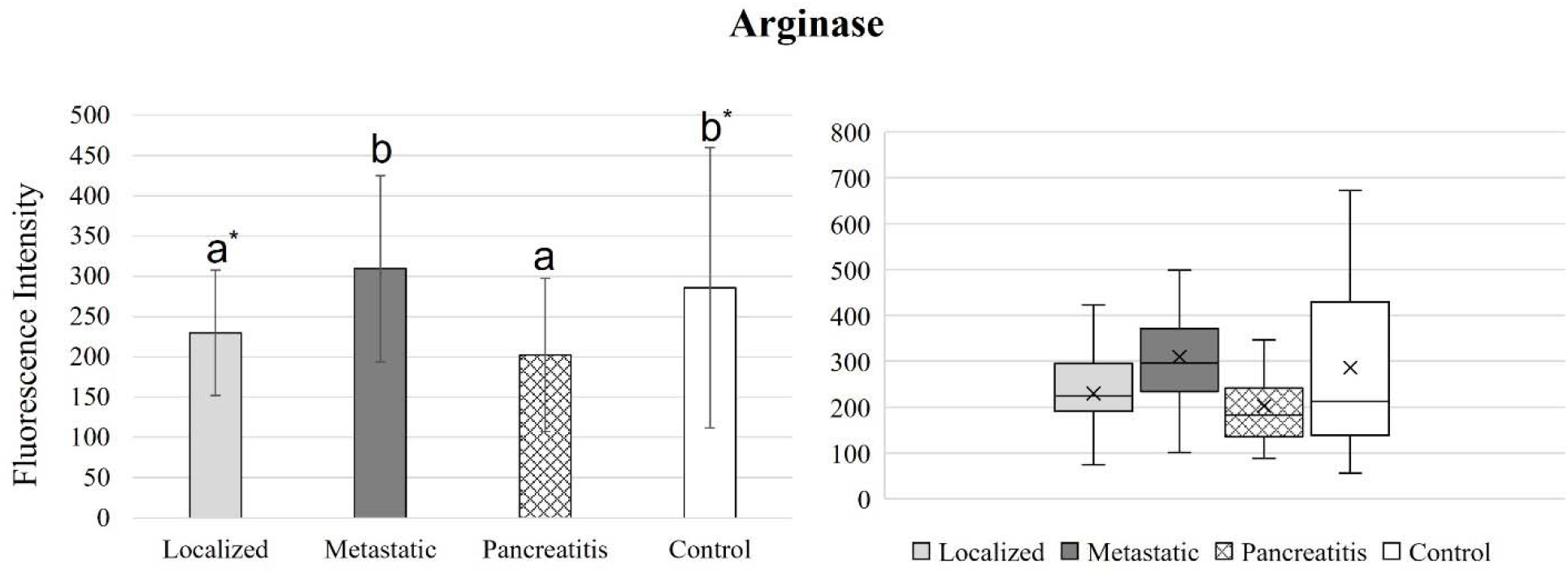
Bar graphs (showing means and standard deviations on the left) and box plots (showing data range on the right) for arginase. Group sizes: localized pancreatic cancer (n=33), metastatic pancreatic cancer (n=50), pancreatitis (n=26), and apparently healthy control volunteers (n=50). Different letters indicate significant differences found in enzymatic activity between disease groups (p-value ≤ 0.05). *Border line significant different (p-value ≤ 0.1 and ≥ 0.05).

**Figure 5.**
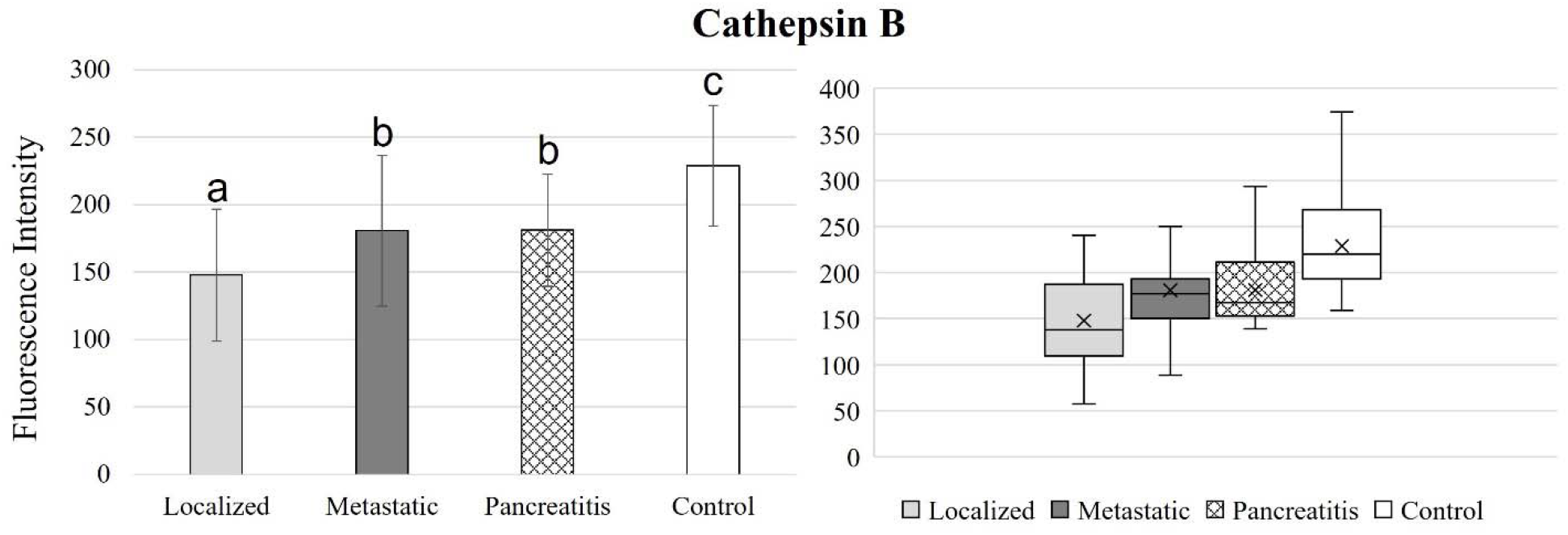
Bar graphs (showing means and standard deviations on the left) and box plots (showing data range on the right) for cathepsin B. Group sizes: localized pancreatic cancer (n=33), metastatic pancreatic cancer (n=50), pancreatitis (n=26), and apparently healthy control volunteers (n=50). Different letters indicate significant differences found in enzymatic activity between disease groups (p-value ≤ 0.05).

**Figure 6.**
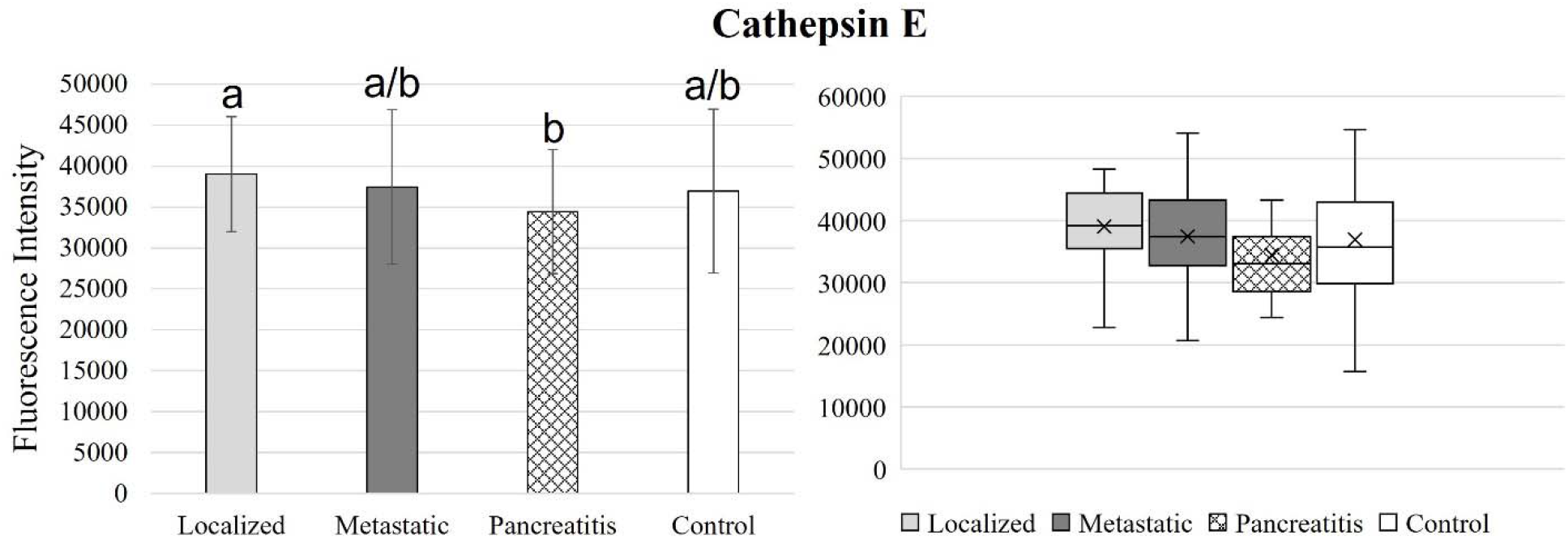
Bar graphs (showing means and standard deviations on the left) and box plots (showing data range on the right) for cathepsin E. Group sizes: localized pancreatic cancer (n=33), metastatic pancreatic cancer (n=50), pancreatitis (n=26), and apparently healthy control volunteers (n=50). Different letters indicate significant differences found in enzymatic activity between disease groups (p-value ≤ 0.05). a/b indicates that group had similar activity with two other groups that were different from each other.

**Figure 7.**
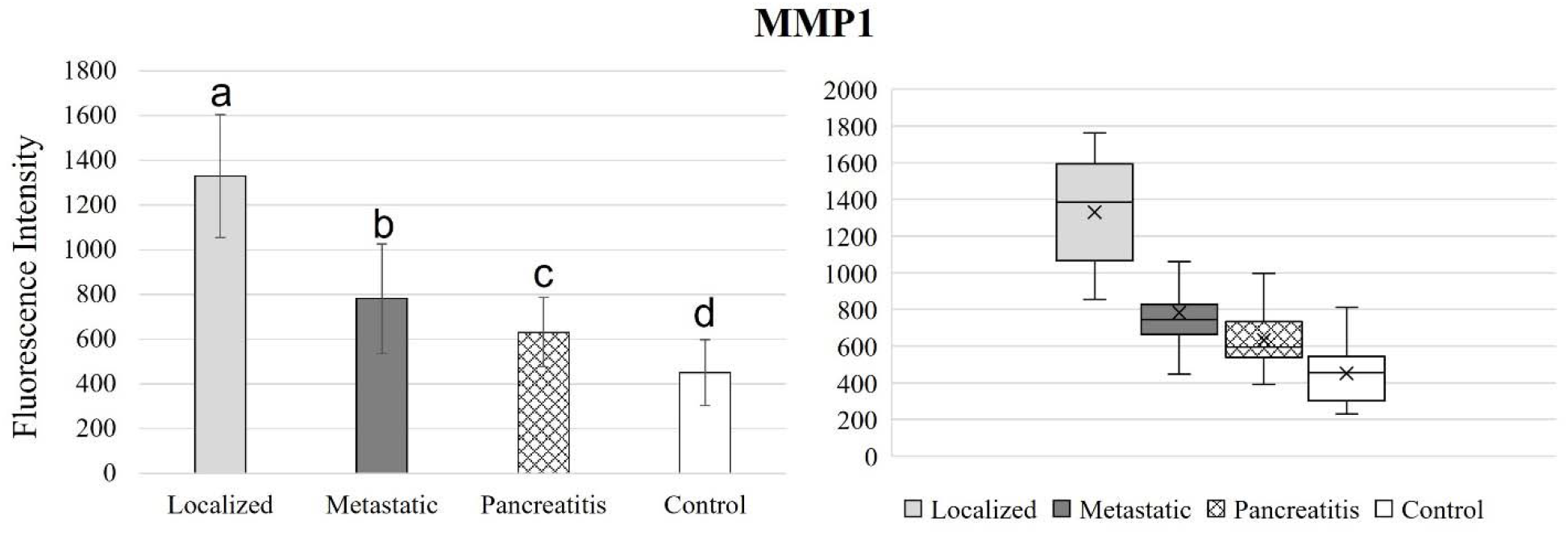
Bar graphs (showing means and standard deviations on the left) and box plots (showing data range on the right) for MMP1. Group sizes: localized pancreatic cancer (n=33), metastatic pancreatic cancer (n=50), pancreatitis (n=26), and apparently healthy control volunteers (n=50). Different letters indicate significant differences found in enzymatic activity between disease groups (p-value ≤ 0.05).

**Figure 8.**
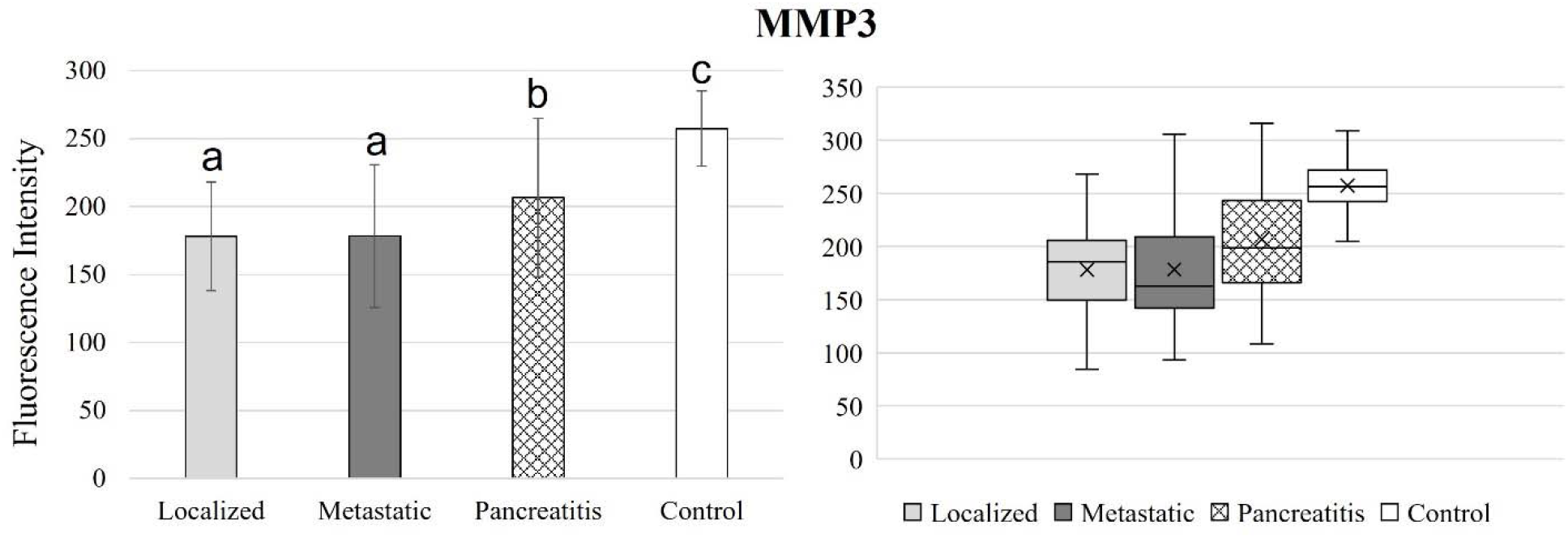
Bar graphs (showing means and standard deviations on the left) and box plots (showing data range on the right) for MMP3. Group sizes: localized pancreatic cancer (n=33), metastatic pancreatic cancer (n=50), pancreatitis (n=26), and apparently healthy control volunteers (n=50). Different letters indicate significant differences found in enzymatic activity between disease groups (p-value ≤ 0.05).

**Figure 9.**
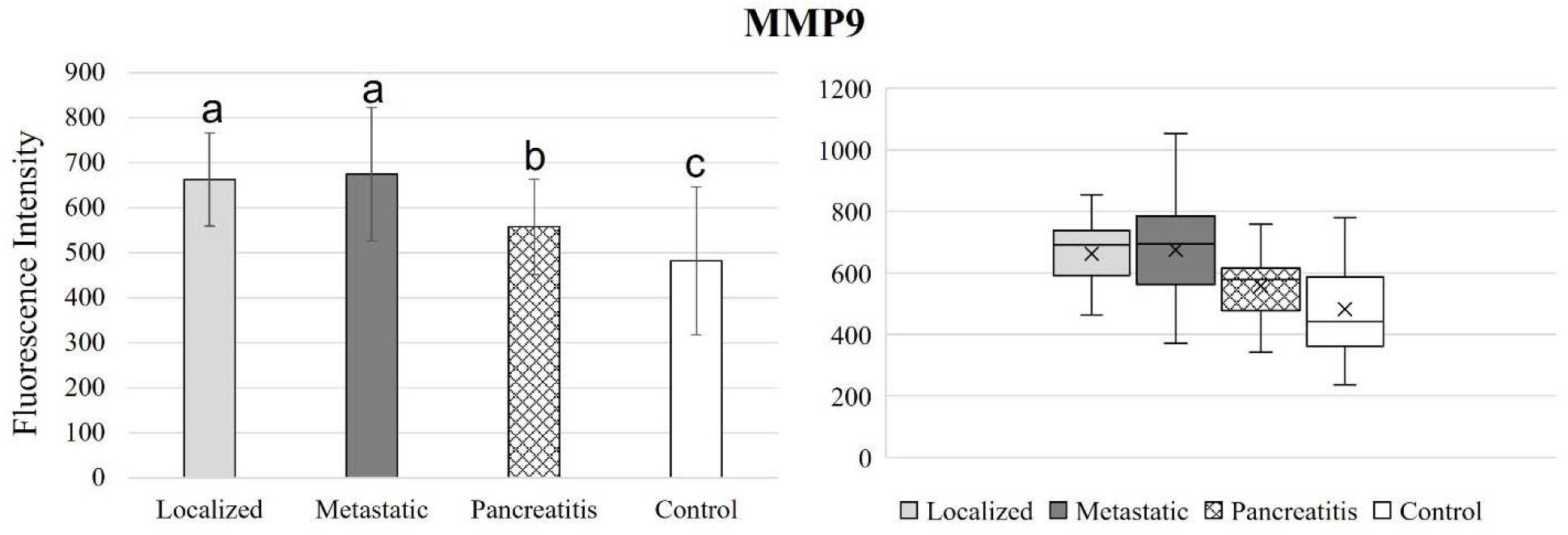
Bar graphs (showing means and standard deviations on the left) and box plots (showing data range on the right) for MMP9. Group sizes: localized pancreatic cancer (n=33), metastatic pancreatic cancer (n=50), pancreatitis (n=26), and apparently healthy control volunteers (n=50). Different letters indicate significant differences found in enzymatic activity between disease groups (p-value ≤ 0.05).

**Figure 10.**
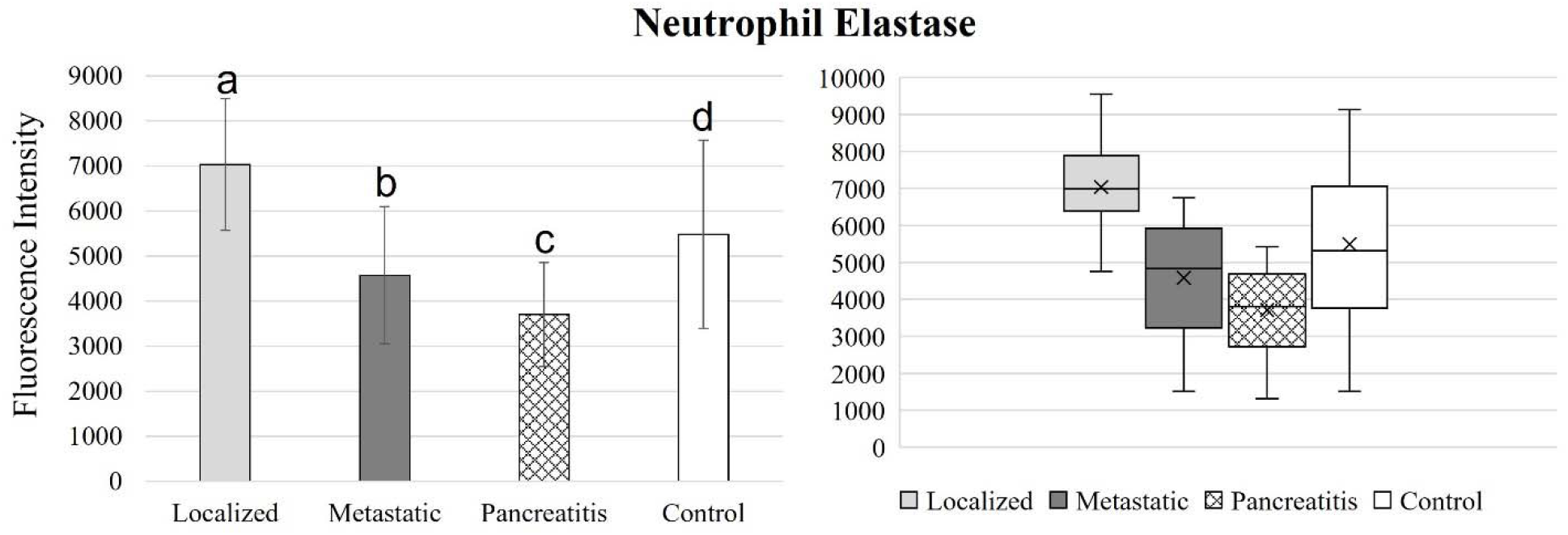
Bar graphs (showing means and standard deviations on the left) and box plots (showing data range on the right) for MMP9. Group sizes: localized pancreatic cancer (n=33), metastatic pancreatic cancer (n=50), pancreatitis (n=26), and apparently healthy control volunteers (n=50). Different letters indicate significant differences found in enzymatic activity between disease groups (p-value ≤ 0.05).

**Figure 11.**
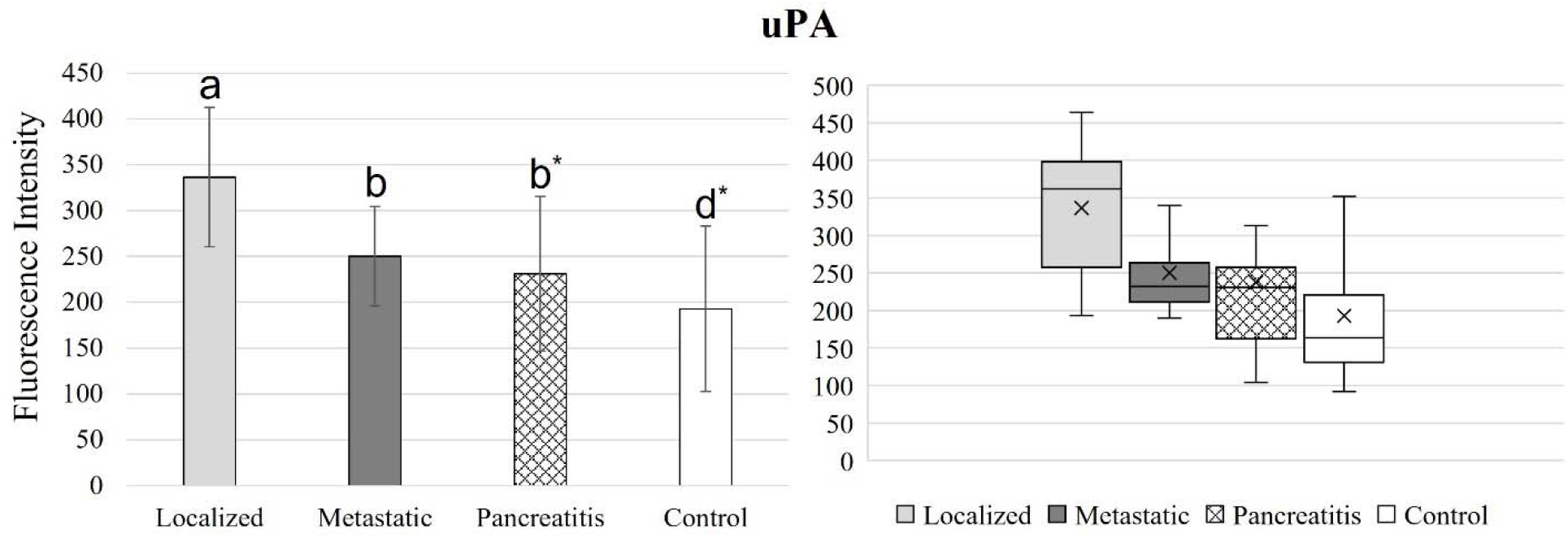
Bar graphs (showing means and standard deviations on the left) and box plots (showing data range on the right) for uPA. Group sizes: localized pancreatic cancer (n=33), metastatic pancreatic cancer (n=50), pancreatitis (n=26), and apparently healthy control volunteers (n=50). Different letters indicate significant differences found in enzymatic activity between disease groups (p-value ≤ 0.05). *Border line significant different (p-value ≤ 0.1 and ≥ 0.05).

Enzymatic activity for cathepsin B was significantly downregulated for all 3 disease groups (localized, metastatic, and pancreatitis) compared to the healthy control group, which indicated that cathepsin B can successfully detect both pancreatic cancer and pancreatitis. While cathepsin B was able to significantly distinguish between localized and metastatic pancreatic cancer as well as localized pancreatic cancer and pancreatitis, it was not able to distinguish between metastatic pancreatic cancer and pancreatitis. Cathepsin B has a strong potential to be used as biomarker for an early detection of pancreatic cancer.

Cathepsin E activity levels remained similar for all disease groups, except for localized pancreatic cancer and pancreatitis. This study indicated that cathepsin E nanobiosensor is not a suitable biomarker in serum for either pancreatic cancer or pancreatitis detection.

MMP1 activity was significantly different for all 4 groups tested, which indicated MMP1 to be a promising candidate for both pancreatic cancer and pancreatitis detection as well as pancreatic cancer staging. Enzymatic activity of MMP1 was downregulated for the healthy control group and increased steadily for pancreatitis, metastatic pancreatic cancer, and localized pancreatic cancer, respectively.

Enzymatic activity decreased steadily from the healthy control group to pancreatitis, metastatic pancreatic cancer, and localized pancreatic cancer, respectively. Even though MMP3 was able to detect both pancreatic cancer and pancreatitis and significantly distinguish between both localized and metastatic pancreatic cancer against pancreatitis, it failed to find a significant difference between localized and metastatic pancreatic cancer groups. Therefore, MMP3 is a good candidate for pancreatic cancer detection but not for pancreatic cancer staging.

While MMP9 activity performed opposite to MMP3 activity (increased in activity level from the healthy control to pancreatitis, metastatic cancer, and localized pancreatic cancer, respectively), the significant differences between groups tested was similar. MMP9 detected both localized and metastatic pancreatic cancer as well as pancreatitis, but it was not able to significantly distinguish between localized metastatic pancreatic cancer.

Enzymatic activity of neutrophil elastase decreased steadily for localized pancreatic cancer, metastatic pancreatic cancer, and pancreatitis, respectively. The differences in neutrophil elastase activity behaved similarly to MMP1 activity, where enzymatic activity was significantly different for all four groups tested. These results indicated that MMP1 and neutrophil elastase biomarkers are ideal candidates for both pancreatic cancer and pancreatitis detection and distinction, as well as pancreatic cancer staging.

In comparison to MMP1 activity, uPA activity increased steadily for pancreatitis, metastatic pancreatic cancer, and localized pancreatic cancer, respectively. uPA is showing the lowest activity in the healthy control group. However, unlike MMP1, uPA activity levels were not significantly different for all four groups tested.

### Decision Framework Evaluation

The evaluation of the proposed decision framework is performed by training a series of decision models considering several combinations of binary classifiers at each hierarchical level indicated in Figure 2. The following classification methods are considered for individual binary classifiers: (i) Gaussian Naïve Bayes (GNB)^33^, (ii) Decision Tree (DT)^34^, (iii) Support Vector Machine (SVM)^35^, (iv) k-Nearest Neighbors (kNN)^36^, (v) Random Forest Classifier (RFC)^34,36^ and (vi) Logistic Regression (LR)^37^. The selected combinations of classification methods exhibiting an overall accuracy score of more than 90% are enlisted in Table 3.

**Table 3:** Combinations of classification methods exhibiting an overall accuracy score of more than 90%.

| Sr. No. | Classification Method |  |  | Correct Predictions | Accuracy Score |
| --- | --- | --- | --- | --- | --- |
|  | Step 1 | Step 2 | Step 3 |  |  |
| 1 | DT | GNB | kNN | 145/159 | 91.19% |
| 2 | SVM | kNN | DT | 144/159 | 90.57% |
| 3 | kNN | GNB | kNN | 144/159 | 90.57% |
| 4 | kNN | DT | DT | 146/159 | 91.82% |
| 5 | kNN | DT | GNB | 148/159 | 93.08% |
| 6 | kNN | DT | RFC | 150/159 | 94.34% |
| 7 | kNN | kNN | RFC | 154/159 | 96.85% |
| 8 | kNN | RFC | DT | 146/159 | 91.82% |
| 9 | RFC | DT | kNN | 149/159 | 93.71% |
| 10 | RFC | DT | RFC | 146/159 | 91.82% |
| 11 | RFC | kNN | kNN | 150/159 | 94.34% |
| 12 | RFC | GNB | SVM | 144/159 | 90.57% |

The training sets were built as elucidated in Methods, and decision models were evaluated over all instances in the dataset. It can be observed in Table 3 that the best performance is obtained using kNN for binary classification in the first two hierarchical levels and RFC in the last step. The corresponding accuracy score is 96.85% and the confusion matrix is provided in Table 4. On the contrary, the traditional multi-class classification approaches yield a maximum classification accuracy of 76.4%, as exhibited in Table 5. This illustrates that the proposed HDS framework significantly outperforms standard multi-class classification approaches for early-stage detection of pancreatic cancer.

**Table 4:** Confusion Matrix – HDS (Step 1: kNN; Step 2: kNN; Step 3: RFC).

| True Class | Predicted Class |  |  |  |  |
| --- | --- | --- | --- | --- | --- |
|  |  | Healthy | Pancreatitis | Localized | Metastatic |
|  | Healthy | 49 | 0 | 0 | 1 |
|  | Pancreatitis | 1 | 25 | 0 | 0 |
|  | Localized | 0 | 0 | 31 | 2 |
|  | Metastatic | 0 | 1 | 0 | 49 |

**Table 5:** Performance obtained using traditional multi-class classification approaches.

| Sr. No. | Classification Method | Accuracy Score |
| --- | --- | --- |
| 1 | GNB | 74.8% |
| 2 | DT | 70.9% |
| 3 | SVM | 76.4% |
| 4 | kNN | 72.4% |
| 5 | RFC | 74.8% |
| 6 | LR | 71.2% |

Results from the use of the SDS framework are shown in Figures 12 and 13. Specifically, Figure 12 illustrates an example scenario with confidence values associated with the four classes showing the basis for a correct prediction. The sample presented in Figure 12 is correctly classified as “Healthy”. It can be observed that there is a significant difference between the probability of this sample belonging to “Healthy” class and that of the other three classes. This depicts a HIGH confidence situation. In such a case, the clinician can safely assume sufficient confidence on the prediction of the decision model and need not prescribe any further diagnostic tests.

**Figure 12.**
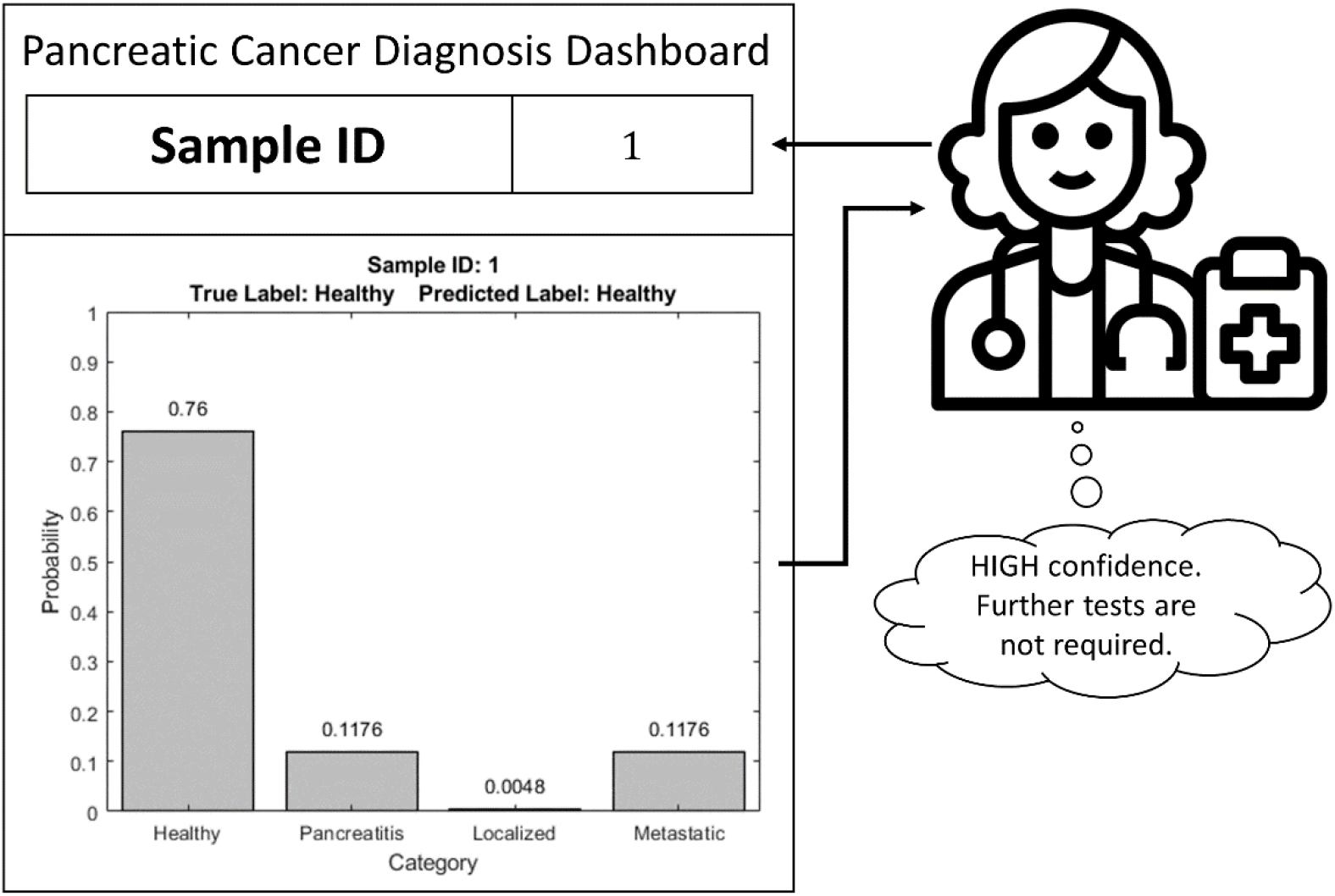
Example of correct prediction by soft hierarchical decision structure.

**Figure 13.**
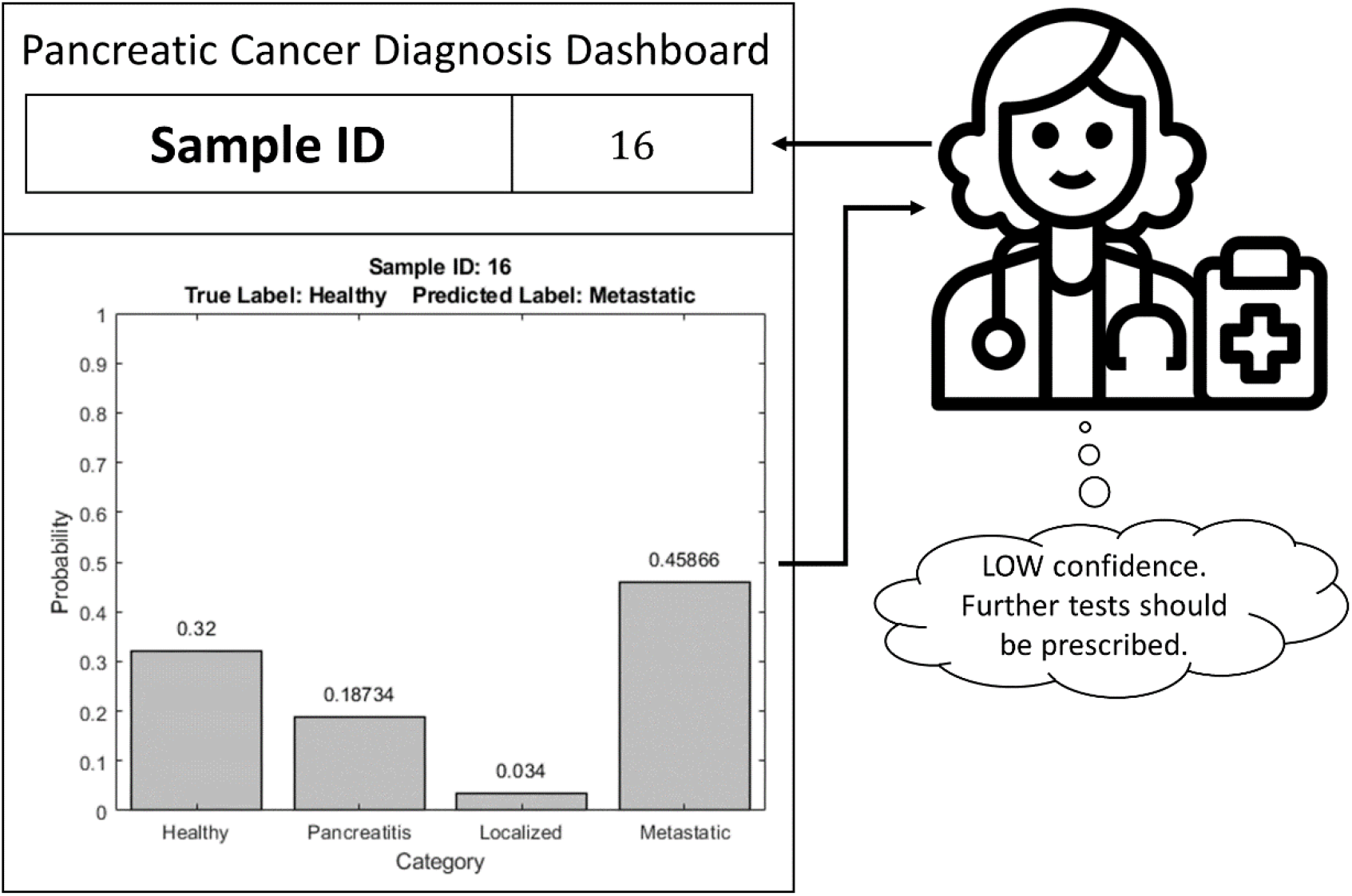
Example of incorrect prediction by soft hierarchical decision structure.

Figure 13 highlights an example wherein the class probabilities do not show a clear favorite, thereby leading to an incorrect prediction. Here, the sample belongs to the “Healthy” class but is misclassified as “Metastatic”. Moreover, it can be observed that the difference in probabilities of “Healthy” and “Metastatic” classes is not that significant as in the previous case, and hence, this is a LOW confidence situation. The presentation of both the decision and class probabilities help clinicians recognize instances where there is a lack of confidence in predictions of the decision model and nudge them to possibly prescribe further tests for better diagnosis. In this manner, the SDS frames the output of the decision model to make it more “interpretable” to the clinicians and possibly use it as a decision support tool while performing diagnosis.

## DISCUSSION

This research with a larger sample size validates our earlier smaller study in 2018 where we had principally demonstrated that pancreatic cancer has a significant protease/arginase signature^13^. These earlier results were confirmed by this study: six out of the eight nanobiosensors tested demonstrated significant differences between both pancreatic cancer groups and the healthy control group, which indicated that these are promising candidates for pancreatic cancer detection (cathepsin B, MMP1, -3, -9, neutrophil elastase, and uPA)^13^. When comparing the pancreatitis against the healthy control group, arginase, cathepsin B, MMP1, -3, -9, and neutrophil, elastase demonstrated to have significant differences in measured fluorescence signal. An ideal panel of biomarkers not only should be able to detect and stage pancreatic cancer, but it should also distinguish between pancreatic cancer and pancreatitis to avoid misleading diagnosis. To optimize the detection probability of especially early (ductal) pancreatic cancer, we have performed in depth data analysis using hierarchical decision structure.

Table 6 summarizes the p-values obtained after comparing activity between each group tested. Columns highlighted in gray represent each disease group compared against the healthy control group, or disease detection. Gray highlighted areas indicate significant differences in fluorescence signal measures (p-value ≤ 0.05). Underlined values indicate borderline significant differences in measured fluorescence signal (0.05 ≤ p-value ≥ 1).

**Table 6.** Significance table between each nanobiosensor and disease group tested

| P-values | Localized<br>vs<br>Control | Metastatic<br>vs<br>Control | Pancreatitis<br>vs<br>Control | Localized<br>vs<br>Metastatic | Localized<br>vs<br>Pancreatitis | Metastatic<br>vs<br>Pancreatitis |
| --- | --- | --- | --- | --- | --- | --- |
| Arginase | <u>0.05082</u> | 0.42605 | 0.00852 | 0.00034 | 0.23682 | 0.00006 |
| Cathepsin B | 1.33E-10 | 7.06E-06 | 1.44E-05 | 0.00584 | 0.00662 | 0.97528 |
| Cathepsin E | **0.27327** | **0.80162** | **0.22787** | 0.38752 | 0.02143 | 0.13966 |
| MMP1 | 6.66E-21 | 3.08E-12 | 1.09E-05 | 2.03E-13 | 4.31E-17 | 0.00170 |
| MMP3 | 1.39E-13 | 2.51E-14 | 0.00021 | 0.98700 | 0.03863 | 0.04254 |
| MMP9 | 2.77E-08 | 1.73E-08 | 0.01780 | 0.66200 | 0.00035 | 0.00019 |
| Neutrophil<br>Elastase | 0.00015 | 0.01430 | 8.42E-06 | 2.29E-10 | 8.44E-14 | 0.00701 |
| uPA | 2.55E-11 | 0.00024 | <u>**0.07370**</u> | 6.58E-07 | 8.05E-06 | 0.30435 |

## Conclusion

One of the major drawbacks of the conventional statistical learning-based or AI-based decision-making algorithms is that they try to enforce a strict decision as an output by selecting the best possible option among the available alternatives. The trust and reliability of the corresponding decisions remain questionable, especially in critical applications like medical diagnosis. The proposed SDS framework provides feedback to clinicians in terms of confidence levels corresponding to each of the possible decisions. The corresponding accuracy score of 96.85%, underscores that a new standard of care in early pancreatic cancer diagnosis by means of Liquid Biopsies is within reach. On the contrary, the traditional multi-class classification approaches yield only a maximum classification accuracy of 76.4%. This illustrates that the proposed HDS framework significantly outperforms standard multi-class classification approaches for early-stage detection of pancreatic cancer. Such feedback is more “interpretable” for the clinicians and therefore, the proposed framework can be potentially deployed as a decision support tool for early-stage detection of pancreatic cancers. It should also be noted that protease/arginase sensing in serum for early pancreatic cancer detection currently not only outperforms genetic tests, but it is also significantly less expensive and relies on clinical plate readers that are already installed in virtually every hospital.

## Data Availability

All data produced in the present study are available upon reasonable request to the authors

## Abbreviations

CTS: Cathepsin
Cy5.5L: cyanine 5.5
Fe/Fe_3_O_4_: iron/iron oxide core/shell nanoparticle
HEPES: 2-[4-(2-hydroxyethyl)piperazin-1-yl]ethanesulfonic acid
MMP: Matrix Metalloprotease
TCPP: tetrakis-carboxyphenyl-porphyrin
uPA: urokinase plasminogen activator

## Author Contributions

The authors have made the following significant contributions to the submitted manuscript: Obdulia Covarrubias-Zambrano: NIH Gene Omnibus analysis, selection of proteases, assembly of the Fe/Fe_3_O_4_-nanobiosensors, nanobiosensor validation, protease activity measurements, writing of a partial draft of the manuscript

Deepesh Agarwal: performing statistical data analysis, writing of a partial draft of the manuscript Madumali Kalubowilage: assembly of the Fe/Fe_3_O_4_-nanobiosensors, protease activity measurements,

Sumia Ehsan: peptide synthesis, characterization, and attachment of TCPP, nanobiosensor validation,

Asanka S. Yapa: TCPP and cyanine 5.5 synthesis, attachment of cyanine 5.5 to the Fe/Fe_3_O_4_-nanobiosensors, nanobiosensor calibration

Jose Covarrubias: synthesis and characterization of Fe/Fe_3_O_4_-nanobiosensors Anup Kasi: clinical guidance, data analysis, writing of a part of the document

Balasubramaniam Natarajan: design of the statistical analysis, writing of parts of the manuscript Stefan H. Bossmann: nanobiosensor design, experimental planning, data analysis, final version of the manuscript.

## Data Availability

The authors declare that the data supporting the findings of this study is available within the paper and its supplementary information files.

